# Covid-19 vs BCG Universal Immunization: Statistical Significance at Six Months of Exposure

**DOI:** 10.1101/2020.09.06.20189423

**Authors:** Serge Dolgikh

## Abstract

With a time-adjusted dataset of Covid-19 statistical data by reporting jurisdiction at the time point of six months after the local epidemics landfall we perform a statistical analysis of the significance of the correlation hypothesis between universal BCG immunization and milder Covid-19 scenarios proposed in the earlier studies. With the data accumulated to date the statistical significance of the BCG immunization correlation hypothesis is evaluated both qualitatively and quantitatively with the conclusion that it has achieved a significant level of confidence. The conclusions of this research can be used in public policy as well as the rationale to investigate the nature and working of a potential broad immunity mechanism associated with an early-age BCG exposure.

## 1 Introduction

A possible correlation between the impact of Covid-19 pandemics and universal immunization program against tuberculosis with BCG vaccine was proposed in [1] and further investigated in a number of other works [2-5]. In this research we compile and provide an analysis of a dataset of cases with different time of local arrival of the epidemics as defined in [2], adjusted and aligned at the time point of approximately six months after the first local exposure.

The intent of this work was to analyze publicly available Covid-19 epidemiological data by reporting national and subnational jurisdictions with respect to the hypothesized induced immunity population-scale protection resulting from a universal BCG vaccination policy (UBIP), current or previous, and attempt both qualitative and quantitative analysis of the hypothesis of correlation between a current or previous UBIP in the jurisdiction and a milder scenario of Covid-19 epidemics; to verify the assumptions, results and conclusions of the earlier studies [1,2,4-6] with a specific objective to determine, in a quantitative analysis, the constraints and confidence of the correlation and null hypotheses.

### 1.1 Terminology

Timing considerations can be critical in the analysis of the development of an epidemiological scenario. For this reason, an effort was made to ensure that the data analyzed was recorded at a similar phase in the development of the epidemics in the jurisdiction. To clarify and emphasize the need for synchronization of epidemiological data, the zero time of the start of the global Covid-19 pandemics was defined in [2] as 31.12.2020: Along with the global Time Zero was defined local Time Zero point (LTZ) indicating the time of arrival of the epidemics in the given locality. It can be sensibly defined as the date of the first confirmed case in the area.

The impact of the epidemics was measured by Covid-19 caused mortality per 1 Million capita in a reporting jurisdiction as a function of time:

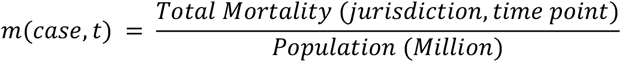

It is believed that this parameter can be a more current and accurate measure of the epidemiological impact than the number of cases that strongly depends on the testing practice, on the assumption that policies and protocols in the selected administration allow reasonably accurate identification of cause and reporting.

Evidently, as defined, the impact of the epidemics in a jurisdiction *M(t)* would be a function of the jurisdiction factors *F*, including demographics, geographical distribution, prosperity, social customs and traditions, lifestyle, public administration and epidemiological policy including records of universal immunization programs and not in the least, time. As the experience of the earlier period shows, the considerations of timing can be, in our view of critical importance for the correctness of the conclusions of the analysis. For that reason, an attention is given to comparing impacts relative to the time of the first exposure by using time-adjusted data at the same local time point in the development of the epidemics.

Throughout this work two related measures of the epidemiological impact will be used as well: relative impact *m*_*R*_*(t)* measured as a ratio of the impact recorded in the locality to the world’s highest value (at the time of evaluation); and the impact expressed in logarithmic scale *m*_*L*_*(t):*

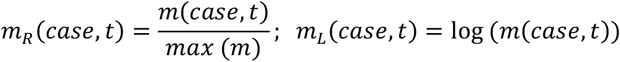

At the time of this study, close to six months after the arrival to jurisdictions in the study, it is expected that the epidemics has the time to develop, and the numbers and relationships to have a more stable nature.

## 2 Methodology

We use qualitative methods such as case comparison, trend analysis and quantitative ones such evaluation of statistical parameters to analyze trends in development of the epidemiological situation across monitored jurisdictions with the intent to evaluate the significance of the correlation hypothesis between the impact of Covid-19 epidemics and a record of universal BCG immunization.

More specifically, the hypothesis that will be evaluated in the study is that of the early age induced broad population-wide protection effect against viral infections including Covid-19 at a certain population-scale level (importantly to note, not necessarily a complete personal protection as in the case of regular vaccinations; and provided the immunization program was implemented consistently, with adequate quality and without significant interruptions). The hypothesis was proposed in [2] based on the analysis of early statistical data and the earlier results in immunology suggesting a possibility of such link [7-9]. To the best of our knowledge, the exact mechanism of such a protection in the immune system has yet to be determined and will be addressed elsewhere; however, a logical possibility that such an induced general immunity protection may have an influence on the epidemiological scenario via providing some level of population-wide mitigation of negative Covid-19 impacts in our view, can be tested with the publicly available epidemiological data.

To evaluate the statistical significance of the correlation hypothesis, we created groups of cases based on UBIP record. This selection is blind based only on the factors of UBIP policy for the jurisdiction irrespective of the current Covid-19 impact statistics. Under the null hypothesis, the cases in different groups should have no significant correlation to the outcomes and therefore, have similar distribution; whereas detecting a significant variation in the outcomes between groups can place constraints on the null hypothesis. The detailed method of evaluation of the statistical significance of the correlation hypothesis is described in Section 5.

## 3 Data

A time-adjusted selected jurisdictions dataset was compiled from public sources with data of Wave 1 and Wave 2 [2] cases adjusted by the time of first exposure to Covid-19. Specifically, the dataset is comprised of the Wave 1 cases as of ∼ TZ + 7 months and Wave 2 cases as of, approximately, TZ + 8 months, i.e. with approximately the same local exposure of six months. It is expected that by this stage, the epidemiological situation has developed to an expressed state in the analyzed jurisdictions.

A number of criteria were applied to selecting the cases in the dataset to minimize the uncertainty factors due to vast variation of conditions among the reporting jurisdictions worldwide:

1. A reasonable expectation of the accuracy, consistency and timeliness of the reporting from the national public health administration;
2. A reasonable level of exposure to Covid-19, e.g. certain minimum number of reported cases and / or impact;
3. A compatible level of social development and specifically, certain minimum standard of public health administration with respect to universal policy administration including importantly, the quality and the coverage. The aim of these criteria is to reduce the uncertainty related to the quality of administration of mass public health policy even when such has been declared.

Categories of cases by the reported epidemiological impact were defined based on the logarithmic scale as follows:

*Very Low* (VL), relative mortality per capita, *m*_*R*_. in the region of 0.001

*Low* (L): *m*_*R*_ ∼ 0.01

*Medium*: (M): *m*_*R*_ ∼ 0.1

*High* (H): Relative m.p.c. higher than 0.2.

While the data for a number of smaller jurisdictions, with population under 5 million was recorded in the dataset, they were not included in the statistical analysis due to higher probability of fluctuations related to unpredictable patterns of cluster development.

BCG universal vaccination record is described by the following bands as defined in [10] and summarized, with modifications and additions, in Table 1:

**Table 1.** BCG universal immunization levels.

| Level | Description |
| --- | --- |
| **A** | Has a current, universal or near-universal BCG vaccination program |
| **A2** | Has a current UBIP with some limitations or qualifications (such as a late start; inconsistencies in application practice, possible significant interruptions due to social factors and other) |
| **B** | Had UBIP in the past covering significant part of population (> 50%) |
| **B2; B3** | UBIP was offered for a limited time interval or specific groups; UBIP practice inconsistent with the hypothesis of early age induced immunity protection for example, delivered at an older age |
| **C** | Never had a universal BCG immunization program |

### Further notes and qualifications

1. Consistency and reliability of data reported by the national, regional and local health administrations.
2. Alignment in the time of reporting may be an issue due to reporting practices of jurisdictions.
3. Availability, consistency and reliability of historical data and statistics on the administration of immunization programs in the national, regional and so on, jurisdictions can be an issue.

The case dataset can be found in the Appendix. Sources: [11-19] and others.

## 4 Results

### 4.1 Regional Variation Analysis

In certain jurisdictions significant regional variability in administration of BCG vaccination can be noted, providing further information relevant to the correlation hypothesis. In the analyzed cases many social parameters, such as living standard, age distribution, social traditions and practices are similar that can be expected to exclude or mitigate the influence of such factors and provide ground for a more confident conclusions of the correlation analysis.

#### Northern Europe

Adjusted to the same time of local exposure, the four cases of Northern Europe show strong correlation between Covid-19 impact and the time of cessation of BCG UIP (Table 2). These countries share similar levels of prosperity, lifestyle and traditions, climate that allows to eliminate many potential influencing factors.

**Table 2.** Covid-19 impact and cessation of UBIP, Northern Europe, at LTZ+6m.

| Case | Sweden | Denmark | Norway | Finland |
| --- | --- | --- | --- | --- |
| M.p.c. | 568.6 | 111.8 | 48.9 | 60.2 |
| UBIP cessation | 1975 | 1986 | 1995 | 2006 |

A similar pattern can be seen in the cases of Portugal (Group A) and Spain (Group B2), with the relative impact, at the time, of 0.064 and 0.22, respectively. These cases provide anecdotal but consistent over the period of four months [2,6] and clear support for the correlation hypothesis.

### 4.2 Cessation of UBIP vs. Covid-19 Impact

In the group B, where a BCG immunization program existed but was ceased earlier, a strong correlation can be observed between the time of cessation of the UBIP and the severity of Covid-19 impact as shown in the diagram of Fig.1 (the data is time adjusted to LTZ + 6 m):

**Fig 1.**
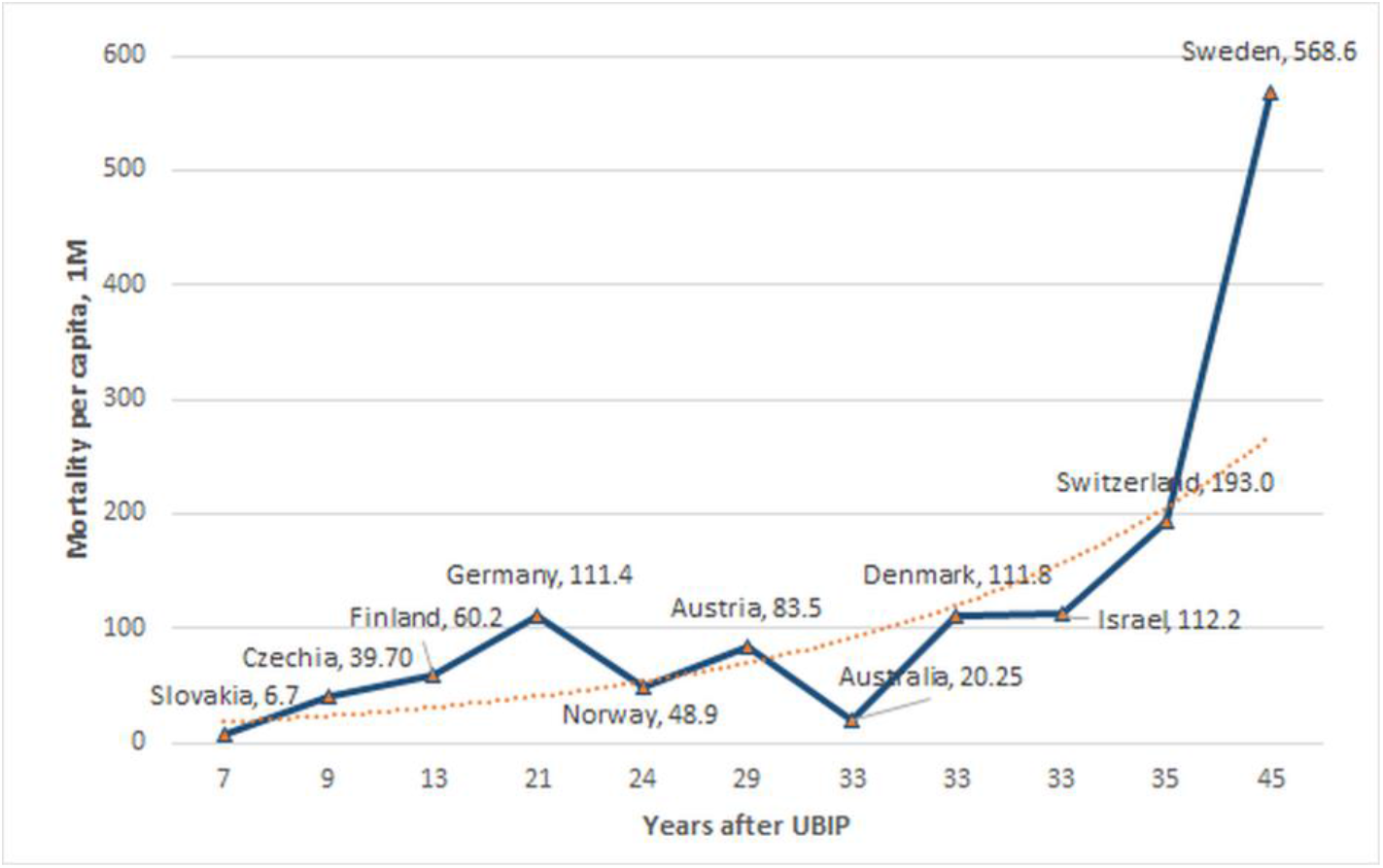
Epidemiological impact vs. time past UBIP.

It can be pointed out that most of the cases in the analysis have similar factors of demographics, levels of prosperity and overall quality of public health administration with the trend of correlation between the time of cessation of UBIP and the Covid-19 impact can be observed clearly. The detailed data used in Fig.1 is provided in Table 2, Appendix.

### 4.3 Heavy Onset Cases, BCG Group A

Several cases of rapid onset of Covid-19 disease were reported in countries with a current BCG UIP, including but probably not limited to, the following: Brazil; Mexico, India, South Africa, Russia, Iran and possibly, others. Without going into specific details of each case that can be done in another study, some general observations can be made here.

As was commented previously [6], in the situations where universality and quality of administration of a UBIP policy in the daily practice could not be ascertained, large group of population may remain without protection even with a declared universal policy. In cases where these vulnerable groups would happen to be more exposed to the infection, it is possible to see higher impact of the epidemics. The factors such as levels of poverty; quality and access to public healthcare; a record of prolonged social disorder, wars, economic collapse and similar could have compromised the administration of UIP. Most of the observed rapid onset cases in group A with a current UBIP fall into one of these categories, though certainly a more detailed analysis of those cases is warranted. Without a specific and detailed study of a jurisdiction it is not possible, in our view, to determine how essential and influential these factors can be and in this analysis we will limit ourselves to stating that given the generally available records for most if not all of such exceptions, they may not be found in a contradiction to the correlation hypothesis.

#### Delayed onset

An interesting observation that can be made about the cases in this group is ostensibly significantly delayed time from the first introduction to the onset of the epidemics. Comparing the cases of the first wave in Europe and Far East one can observe the onset period of approximately 1.5 – 2 months (from end of January to March 2020, when the epidemics was in full development in the European jurisdictions) [1,2]. This can be contrasted to the development period of 4 months and over in many cases discussed in this section: Mexico, Brazil, India, South Africa, Russia and other similar cases. Whilst this observation at the time of writing does not have statistical significance, it can be seen as an indirect argument for the correlation hypothesis, as the possibility that the immunized part of the population delays the transmission of the epidemics to the unprotected groups. This hypothesis requires further analysis and will be addressed elsewhere.

### 4.4 Possible Mechanism

The hypothesis of induced early age immunity protection from the exposure to BCG proposed in [2] based on a number of reports pointing at a possible association between early delivery of BCG vaccine and a broad immunity against several conditions [8-9,20]. It is further supported by a study indicating a possible mechanism for increased production of immune cells in infants following vaccination with BCG [7,21].

## 5 Statistical Significance of the Correlation Hypothesis

In this section the statistical significance analysis conducted previously for the time period of LTZ + 3 months is repeated at the time point of six months after the first local exposure to the infection. The analysis is based on evaluation of statistical parameters such as mean and standard deviation of the epidemiological impact between groups of cases blindly selected based on the record of UBIP as described in detail in [6]. The null hypothesis in this case would dictate that immunization should carry no statistical significance for the epidemics impact, and therefore distributions in all of BCG group sample points (A, B, C) as defined above were described by a single distribution with, possibly, time-dependent parameters *μ(t), σ(t*) that can be estimated from the overall dataset.

The basis for the analysis that follows is the observation of a strong disparity between the sample means in groups A and C as first reported in [1,2,6]. Under the null hypothesis, these cases should be treated as the difference between the means of randomly drawn samples of a given size, for which distribution parameters can be estimated with the sample-mean law.

The groups of cases in the analysis were selected based on the record of UBIP under the discussed criteria. The resulting groups were:

**Group A** (current UBIP and equivalent, protected cohort over 30 years of age): Taiwan, Japan, South Korea, Singapore, Slovakia, Poland, Ukraine, Greece, Portugal, Czechia (2010), Finland (2008), Germany (1998), Norway (1995), Austria (1990).

**Group B** (previous UBIP): Australia (c.1985), Israel (1986), Denmark (1987), Switzerland (c.1985), Sweden (1975).

**Group C** (no record of a previous UBIP, and equivalent): Italy, Belgium, Netherlands, United Kingdom (see Notes below), Spain (Notes), USA, Ontario, Quebec (provinces of Canada), California, Florida, New York (states of USA).

For further notes and explanations on the composition of the groups refer to the Appendix.

Under the assumption of the null hypothesis all group samples would be drawn from the same distribution and the rule of sample means dictates that the means of the

samples of groups A – C with the number of samples N_G_ will be distributed with the same mean and a standard deviation σ_*G*_ as:

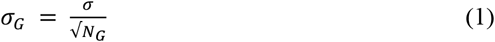

where s is the overall standard deviation of the dataset. From (1) based on the size of each group, one can estimate sample mean standard deviations for the groups A – C samples. For the selected groups of cases, statistical parameters of the epidemiological impact distribution measured in logarithmic mortality per capita *m*_*L*_*(case, t)* were obtained from the dataset (the Appendix) as shown in Table 3.

**Table 3.** Statistical parameters of the epidemiological impact by case group.

| *Sample* | *Number of cases* | *Sample mean* | *Sample STD* | *Sample mean $\sigma$* | *Sample mean deviation, $\sigma$* |
| --- | --- | --- | --- | --- | --- |
| **A** | 14 | 4.50 | 2.46 | 0.75 | -2.76 |
| **B** | 5 | 6.94 | 1.74 | 1.25 | 0.28 |
| **C** | 11 | 9.09 | 0.81 | 0.86 | 2.90 |
| **Overall** | 30 | 6.59 | 2.80 | - | - |

To satisfy the null hypothesis, the means of samples A – C would need to independently, satisfy the normal distribution laws with the same mean *μ*_*S*_ that can be assumed to be equal to the overall dataset mean and *σ*_*G*_ defined by (1). Then following the arguments outlined in [6] one obtains:

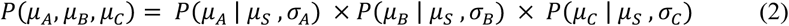

where the first term on the right is the probability of *μ*_*A*_ within the observed range below *μ*_*S*_ with a standard deviation *σ*_*A*_ and the second, similarly, of *μ*_*C*_ within the observed range above *μ*_*S*_ with a standard deviation *σ*_*C*._ Then from (2) the p-value of the null hypothesis can be estimated as:

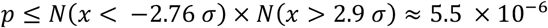

excluding the null hypothesis at a confidence level of at least 10^−5^. The mean of the B-sample was close to the dataset mean *μ*_*s*_ and for that reason did not contribute significantly to the p-value constraint.

This result can be illustrated by a histogram of the epidemiological impact for cases in UBIP groups A and C (Fig.3) in the analysis above.

**Fig 2.**
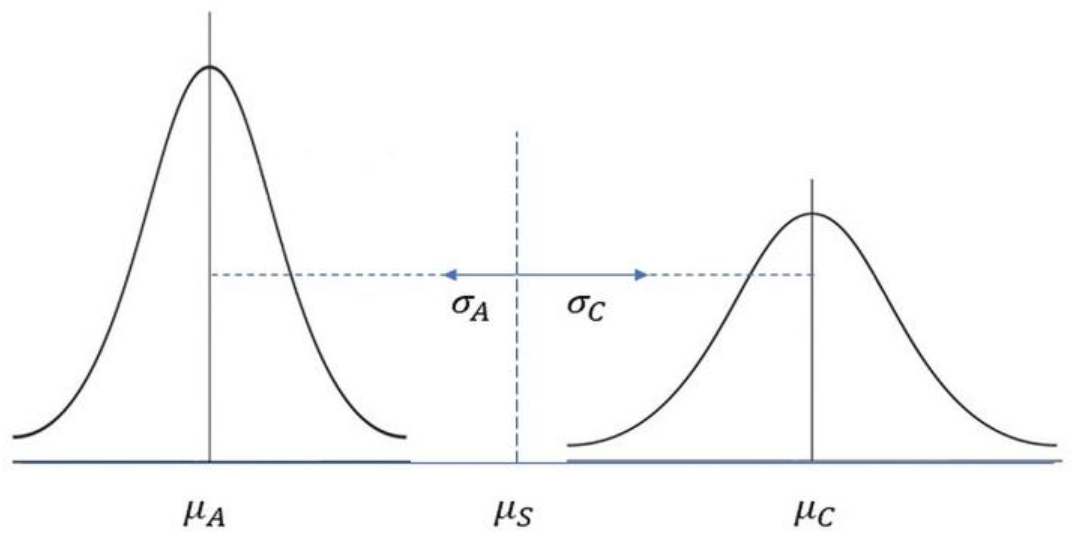
Sample means distributions, UBIP case groups.

**Fig 3.**
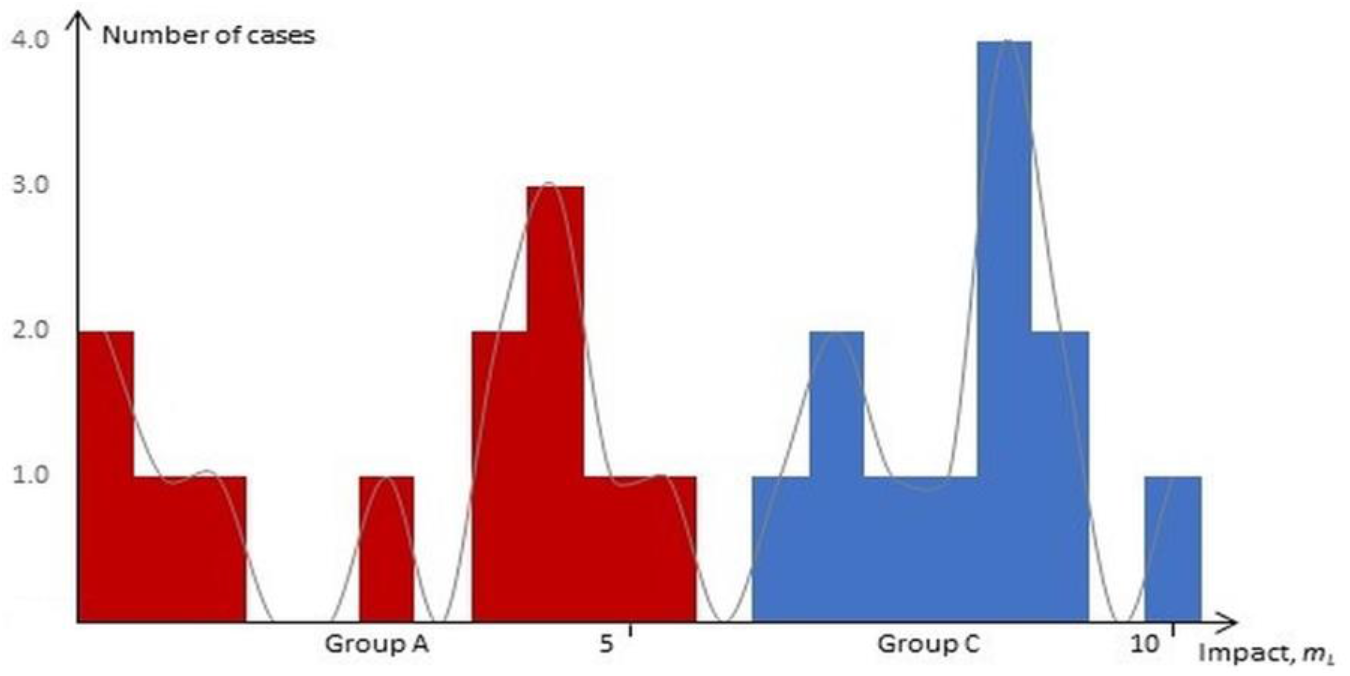
Case distribution histogram, UBIP groups A and C.

The trend of the groups A (red) and C (blue) to the different ends of the impact range (in logarithmic mortality per capita) can be seen clearly, supporting the results of the statistical significance analysis.

To summarize the results of this section, if the group samples, selected blindly according to the record of UBIP had no correlation with the epidemiological impact and therefore, considered as independent random samples under the null hypothesis, repeated observations of sample means as far apart as in the analyzed groups would lead to strong constraints on the p-value of the null hypothesis.

## 6 Conclusion

With a time-adjusted dataset of epidemiological statistics for national and subnational jurisdictions at the time point of 6 months after the first exposure statistical significance of the correlation hypothesis between the record of universal BCG vaccine immunization performed at birth or early infancy and milder Covid-19 epidemiological scenario in the jurisdiction. The results of several qualitative observations Sections 4.1, 4.2 consistently point at a correlation between these factors.

In addition to convincing, in our view, qualitative arguments in support of the correlation hypothesis, a statistical analysis of the correlation between UBIP record and current epidemiological impact in Section 5 confirms statistical significance of the correlation hypothesis with a confidence of at least 0.0001. The result is consistent with the analysis of statistical significance at the time of exposure of 3 months [6] and increases the confidence in the overall conclusion because under the null hypothesis samples taken at different time intervals should be considered as independent as well.

The findings of this research are in agreement with the earlier studies [1-6] supporting the correlation hypothesis and in our view, provide a strong rationale for further research into possible mechanisms for such a broad induced protection with the potential of developing effective methods of long-term immunity against a broad range of diseases.

It is hoped that time-adjusted datasets compiled in this work as the early observations obtained with it can be useful to other researchers in the field looking for effective approaches to understanding and eventually, effectively managing and controlling this and similar infectious diseases in the future.

## Data Availability

Dataset is available and provided

https://news.google.com/covid19/map

## Appendix

### 1. Case Dataset

Time-Adjusted Combined Dataset, LTZ + 6 months (updated 03.09.2020)

| *Case* | *Population*<br>*(M)* | *Local*<br>*landfall* | *Mortal-*<br>*ity, per*<br>*IM* | *M.p.c,*<br>*relative* | *UBIP type,*<br>*cessation* |
| --- | --- | --- | --- | --- | --- |
| **Very Low** |  |  |  |  |  |
| Taiwan | 23.8 | 21.01 | 0.29 | 0.000 | A |
| Japan | 126.8 | 16.01 | 9.31 | 0.003 | A |
| South Korea | 51.5 | 20.01 | 5.88 | 0.002 | A |
| Singapore | 5.6 | 23.01 | 4.64 | 0.002 | A |
| Slovakia | 5.5 | 6.03 | 6.73 | 0.002 | A |
| **Low** |  |  |  |  |  |
| Australia | 24.6 | 25.01 | 20.24 | 0.007 | B, c.1985 |
| Greece | 10.5 | 28.02 | 26.0 | 0.009 | A |
| Czechia | 10.7 | 01.03 | 39.72 | 0.014 | B, 2010 |
| Croatia | 4.1 | 25.02 | 47.32 | 0.017 | A |
| Norway | 5.4 | 26.02 | 48.89 | 0.018 | B, 1995 |
| Poland | 38.0 | 04.03 | 55.05 | 0.020 | A |
| Ukraine | 42.2 | 29.02 | 67.06 | 0.024 | A |
| Kyiv, Ukraine | 3.8 | 16.03 | 60.53 | 0.022 | A |
| Finland | 5.5 | 29.01 | 60.18 | 0.022 | B, 2006 |
| Austria | 8.8 | 25.02 | 83.52 | 0.030 | B, 1990 |
| Albania | 2.9 | 08.03 | 105.71 | 0.038 | A |
| Israel | 8.7 | 21.02 | 112.20 | 0.041 | B, 1986 |
| Germany | 82.8 | 27.01 | 111.43 | 0.041 | B, 1998 |
| Denmark | 5.6 | 27.02 | 111.79 | 0.041 | B, 1986 |
| **Mid** |  |  |  |  |  |
| Portugal | 10.3 | 2.03 | 177.38 | 0.064 | A |
| Switzerland | 8.6 | 25.02 | 193.0 | 0.070 | B, c.1985 |
| California, USA | 39.5 | 26.01 | 236.86 | 0.086 | C |
| Canada | 37.6 | 25.01 | 237.90 | 0.086 | B2 |
| Ontario (Canada) | 14.6 | 25.01 | 190.27 | 0.069 | C |
| Argentina | 45.2 | 3.03 | 203.27 | 0.074 | A |
| South Africa | 57.8 | 5.03 | 248.94 | 0.090 | A |
| Ireland | 4.9 | 29.02 | 362.65 | 0.132 | A |
| Netherlands | 17.2 | 27.02 | 362.50 | 0.132 | C |
| Florida, USA | 21.5 | 1.03 | 541.81 | 0.197 | C |
| United States | 327.2 | 21.01 | 408.38 | 0.175 | C |
| France | 67.0 | 24.01 | 451.72 | 0.164 | B3 (2007) |
| Sweden | 10.1 | 31.01 | 568.61 | 0.207 | B, 1975 |
| Ecuador | 17.6 | 29.02 | 376.08 | 0.137 | B, unknown |
| Mexico | 126.2 | 28.02 | 521.52 | 0.190 | A |

| High |  |  |  |  |  |
| --- | --- | --- | --- | --- | --- |
| Italy | 60.5 | 31.01 | 581.06 | 0.211 | C |
| Spain | 46.7 | 31.01 | 609.10 | 0.221 | B2, 1965-81 |
| Quebec, Canada | 8.5 | 28.02 | 678.12 | 0.246 | B2, 1956-74 |
| United Kingdom | 66.4 | 29.01 | 695.80 | 0.253 | B3 (2005) |
| Brazil | 210 | 25.02 | 590.00 | 0.214 | A2 |
| Chile | 18.1 | 03.03 | 626.74 | 0.280 | A |
| Belgium | 11.4 | 04.02 | 863.60 | 0.314 | C |
| New York (State), USA | 19.4 | 1.03 | 1678.71 | 0.610 | C |
| New York (City), USA | 8.6 | ~15.02 | 2751.16 | **1.00** | C |

### 2. Time-Adjusted Impacts, Jurisdictions with Previous UBIP (updated 03.09.2020)

| *Case* | *Mortality, per IM* | *UBIP Type* | *UBIP Cessation* |
| --- | --- | --- | --- |
| Czechia | 39.7 | B | 2010 |
| Finland | 60.2 | B | 2006 |
| Germany | 111.4 | B | 1998 |
| Norway | 48.9 | B | 1995 |
| Austria | 83.5 | B | 1990 |
| Australia | 20.2 | B | 1987 |
| Denmark | 111.8 | B | 1986 |
| Israel | 112.2 | B | 1986 |
| Switzerland | 193.0 | B | c.1985 |
| Sweden | 568.6 | B | 1975 |
| Quebec (Canada)<sup>1</sup> | 678.1 | B2 | 1956-74 |
| Spain<sup>1</sup> | 609.1 | B2 | 1965-81 |
| United Kingdom<sup>2</sup> | 695.8 | B3 | 2005 |
| France<sup>3</sup> | 451.72 | B3 | 2007 |

<sup>1</sup> Considered no-UBIP equivalent due to a very short duration of UBIP.
<sup>2</sup> Considered no-UBIP equivalent due to immunization practice incompatible with the early age immunity hypothesis.
<sup>3</sup> Inconsistent data about the immunization practice.

### 3. Notes on UBIP Case Groups

1. Equivalent in Group A were the cases with a recent cessation of UBIP, with unprotected under the correlation hypothesis age cohort under 30 years of age.
2. Equivalent in Group C were the cases with a very short or otherwise affected UBIP as further noted below.
3. Group A selection: only cases with a reliable and consistent application of UBIP were selected. The case of Ireland was not added in this group due to documented records of inconsistent policy application across geographical regions [21].
4. The case of Spain was placed in Group C due to a very short duration of UBIP (less than 20 years overall) making it negligible for any hypothesized effect of population-wide protection. The same argument was applied to the case of Quebec, Canada.
5. The case of United Kingdom placed in Group C due to incompatibility of the UBIP administration practice with the early age induced immunity hypothesis as it was administered at a school or early adolescence age [10].
6. Canada, where no UBIP was provided except for a short duration in the province of Quebec [18] was represented by the cases of Ontario and Quebec, with the highest population and epidemiological impact (two cases).
7. Due to large population and high regional variation, United States was represented by three state cases California, Florida, New York, and the overall statistics for the country (four cases).
8. The case of France where UBIP was in place till 2007 was not included in any group due to uncertainty about the administration practice. Sources provide inconsistent information about the age of administration, infancy or school age: “BCG was mandatory for school children between 1950 and 2007” [11,23] indicating a possibility that in the least the practice was not consistent and the decision on the placement in the appropriate group could not be made without further detailed analysis.

